# Biogeneric native and polyethylene glycol-conjugated *E. coli* asparaginases for treating children with acute lymphoblastic leukaemia

**DOI:** 10.1101/2025.10.28.25337165

**Authors:** Neerajana Datta, Bishwaranjan Jana, Srijani Goswami, Annwesha Roy, Samik Samaddar, Subhajit Kundu, Manash Pratim Gogoi, Bony Dasgupta, Trina Dutta, Nandana Das, Vaskar Saha, Jasmeet Sidhu, Shekhar Krishnan

## Abstract

*Escherichia coli* asparaginases (EcASNase) available for the treatment of children with acute lymphoblastic leukaemia (ALL) are largely of suboptimal quality in low-middle income countries (LMIC) and contribute to inferior outcomes. The pharmacokinetics, activity, and immunogenicity of a native (EcASNase) and a PEGylated ASNase (PEG-EcASNase) were analysed. Biogeneric EcASNase 10,000 IU/m^2^ was administered intramuscularly every 72 hours (Cohort 1, 76 patients) or every 48 hours (Cohort 2, 69 patients). Cohort 3 (176 patients) received a PEG-EcASNase biogeneric 1000 IU/m^2^ intramuscularly every 14 days. In Cohort 1, 69% of trough induction samples were suboptimal (<100 IU/L). Cohort 2 achieved a median trough activity >600 IU/L in induction, with no suboptimal activity. In Cohort 3 median trough activity in induction was 467 IU/L with suboptimal activity in 7–16%. Patients receiving one induction PEG-EcASNase dose had eightfold higher odds of suboptimal activity post-induction. Hypersensitivity was higher with EcASNase (12% vs. 5%) and pancreatitis more often with PEG-EcASNAse (5% vs. 1%). Anti-drug antibodies were strongly associated with silent inactivation and hypersensitivity (p=<0.0001). Pharmacological monitoring enabled optimization of EcASNase dosing, detection of silent PEG-EcASNase inactivation and identification of induction dosing intensity as a determinant of sustained activity for generic ASNase products marketed in LMICs.

## Introduction

Asparaginase (ASNase) is a vital drug for the treatment of acute lymphoblastic leukaemia (ALL). Premature ASNase discontinuation due to clinical toxicity (1), or inadequate exposure due to silent inactivation (2), is associated with inferior clinical outcomes. Polyethylene glycol-conjugated formulations of *Escherichia coli*-derived ASNase (Oncaspar, Asparlas; Servier Pharmaceuticals) are the standard of care in higher income countries. In low-middle-income countries (LMICs), the high cost of these innovator products precludes their use. Instead, biogeneric substitutes are marketed in these regions. We and others (3-5) have shown that many biogeneric products are of substandard quality, leading to inadequate therapeutic activity and inferior clinical outcomes (6). To address this problem, we established an ASNase monitoring programme and collaborated with local pharmaceutical companies to systematically evaluate their ASNase products. The primary objective was to identify clinically suitable, locally accessible biogeneric ASNases and establish their optimal dosage and administration schedule. This was accomplished through systematic clinical and pharmacological monitoring within a standardised, risk-stratified treatment protocol. (7) We began by evaluating Leucoginase, a native biogeneric *E. coli* ASNase (EcASNase) from VHB Life Sciences, Mumbai, India; product P6 in our earlier publication (5). We subsequently focused on Hamsyl, an indigenously manufactured biogeneric PEGylated *E. coli* ASNase (PEG-EcASNase) from Gennova Biopharmaceuticals (Pune, India). This PEG-EcASNase biogeneric was demonstrated to be bioequivalent to its innovator counterpart (8) and showed promising therapeutic activity (9). Hamsyl monitoring was implemented through a drug donation and vial-sharing programme and its fewer doses were an advantage during the COVID-19 pandemic restrictions. In this report, we discuss our observations with both Leucoginase and Hamsyl asparaginases and the insights gained from these monitoring studies.

## Patients and Methods

Children and adolescents aged 1-18 years with newly-diagnosed ALL were enrolled in the asparaginase monitoring programme at the Tata Medical Center (Kolkata, India). The study programme was approved by the institutional review board and written informed consent obtained from patients/families. Native or PEG-conjugated *E. coli* ASNase was administered intramuscularly as part of the risk-stratified Indian Collaborative Childhood Leukaemia (ICiCLe-14) treatment protocol.(7) All patients received asparaginase during the induction and delayed intensification treatment phases. In patients with high-risk B cell-precursor ALL and T-lymphoblastic leukaemia/lymphoma, additional ASNase doses were administered during the consolidation (augmented Berlin-Frankfurt-Münster) and interim maintenance (Capizzi methotrexate) treatment blocks. Between March 2018 and March 2020, patients received Leucoginase, a native EcASNase biogeneric (VHB Life Sciences; Mumbai, India) at 10,000 IU/m^2^/dose intramuscularly. This was initially administered every 72 hours (Cohort 1; March 2018 to June 2019) and later every 48 hours (Cohort 2, June 2019 to March 2020). A subsequent cohort received Hamsyl, a PEG-EcASNase biogeneric (Gennova Biopharmaceuticals; Pune, India) administered intramuscularly at 1000 IU/m^2^/dose every 14 days (Cohort 3; August 2020 to March 2023). Details of ASNase treatment, dosing and schedule in the different cohorts and ALL risk groups are presented in 0. Data and Supplementary Table S1. ASNase-associated toxicities, chiefly hypersensitivity and pancreatitis, were defined following the consensus guidelines of the Ponte di Legno toxicity working group (10) and revised Atlanta criteria (11). Hepatotoxicity and thromboembolism were defined using v5·0 of the Common Terminology Criteria for Adverse Events. All cases of asparaginase-associated toxicities were reviewed by clinicians and study investigators.

Plasma asparaginase activity (PAA) was measured using the aspartate-β-hydroxamate / indooxine assay as reported previously (12). Inter-day and intra-day assay parameters for accuracy, precision, and recovery were maintained within ± 10% of nominal values, with a lower limit of quantification (LLOQ) of 10 IU/L. PAA was reported at trough timepoints (72 hours post-dose in Cohort 1; 48 hours post-dose in Cohort 2; 14±2 days post-dose in Cohort 3). Trough PAA values ≥100 IU/L were considered adequate. Trough PAA measurements below 100 IU/L were defined as suboptimal, while measurements below LLOQ were considered inadequate. Silent inactivation was inferred when trough PAA levels were suboptimal or inadequate without accompanying clinical hypersensitivity (13). Leucoginase activity-time course was plotted by measuring PAA in serial samples collected 6, 12, 24, 48 and 72 hours following the first intramuscular dose of Leucoginase. Additional PAA measurements were carried out in patients with suspected ASNase-associated clinical hypersensitivity.

Anti-ASNase IgG antibodies were detected and quantified using a modified indirect enzyme - linked immunosorbent assay (14, 15). Anti-drug antibody testing was carried out serially in the following Cohort 3 patient subsets: (i) patients with clinical hypersensitivity, (ii) patients with persistent inadequate trough PAA (iii) patients with probable silent inactivation, and (iv) a subset with adequate trough PAA levels across treatment phases. Selection criteria for the fourth subset included patients with (a) sustained adequate PAA across all treatment phases, (b) availability of at least one trough time-point sample from induction and post-induction phases to evaluate antibody development in response to repeated dosing, (c) sampling within six months of testing to ensure both suitable sample quality and ready retrievability of stored samples and (d) trough PAA values within 1·5 times the interquartile range of PAA measurements for Cohort 3, to exclude outliers and maintain representative activity profiles. Assay information and cut-off determinations are detailed in the Supplementary Section.

Continuous variables were represented as median with interquartile ranges. Groups with continuous variables were compared using the Mann–Whitney U or Kruskall–Wallis tests as appropriate. Categorical variables were compared using the chi-squared or the Fisher exact tests as appropriate. Factors influencing PAA were evaluated using generalised estimating equations for repeated measures and an exchangeable correlation structure. Two-sided p values were determined for all tests. Analyses were carried out using GraphPad Prism 10·2·2 (GraphPad Software Inc.) and SPSS statistical software (v26·0, IBM Corp), and figures were plotted using GraphPad Prism.

## Results

Between March 2018 and March 2023, 318 consecutive patients with newly-diagnosed ALL treated uniformly on the risk-stratified protocol ICiCle-14 protocol, were enrolled in the institutional asparaginase monitoring programme. Cohort characteristics are summarised in Supplementary Table S2. In Cohort 1, 109 trough time-point samples (81 in induction, 21 post-induction) from 62 patients were analysed. The median trough plasma asparaginase activity (PAA) was 73 IU/L (interquartile range [IQR], 38–122 IU/L) during induction and 134 IU/L (IQR, 74–226 IU/L) post-induction (p=0·0027). Suboptimal trough levels (trough PAA <100 IU/L) were observed in 69% (56/81) of induction samples and 32% (9/28) of post-induction samples (p=0·0008) (Figure 1A, Supplementary Table S3). A time-course study of Leucoginase activity in six Cohort 1 patients (Figure 1B) showed peak activity 12-24 hours after intramuscular administration, sustained above the 100 IU/L threshold for 48 hours before declining to suboptimal levels 72 hours post-dose (mean PAA at 72 hours: 45 IU/L S.E. 16 IU/L). This finding led to a revised 48-hour Leucoginase dosing schedule for Cohort 2. In Cohort 2, 119 trough time-point samples (91 in induction, 28 post-induction) from 57 patients were analysed. The median trough PAA was significantly higher at 617 IU/L (IQR, 314–918 IU/L) during induction and 1161 IU/L (IQR, 639–1651 IU/L) post-induction (p=0.0007). No samples in this cohort had suboptimal activity levels (Figure 1C, Supplementary Table S3). In Cohort 3, 537 trough time-point samples (225 in induction, 312 post-induction) from 167 patients were analysed. The median trough PAA was 467 IU/L (IQR, 317–630) during induction and 633 IU/L (IQR, 403–835) post-induction (p<0·0001). Suboptimal trough levels were observed in 7% (16/225) of induction samples and 16% (49/312) of post-induction samples (p=0·0027) (Figure 1D, Supplementary Table S3).

**Figure 1.**
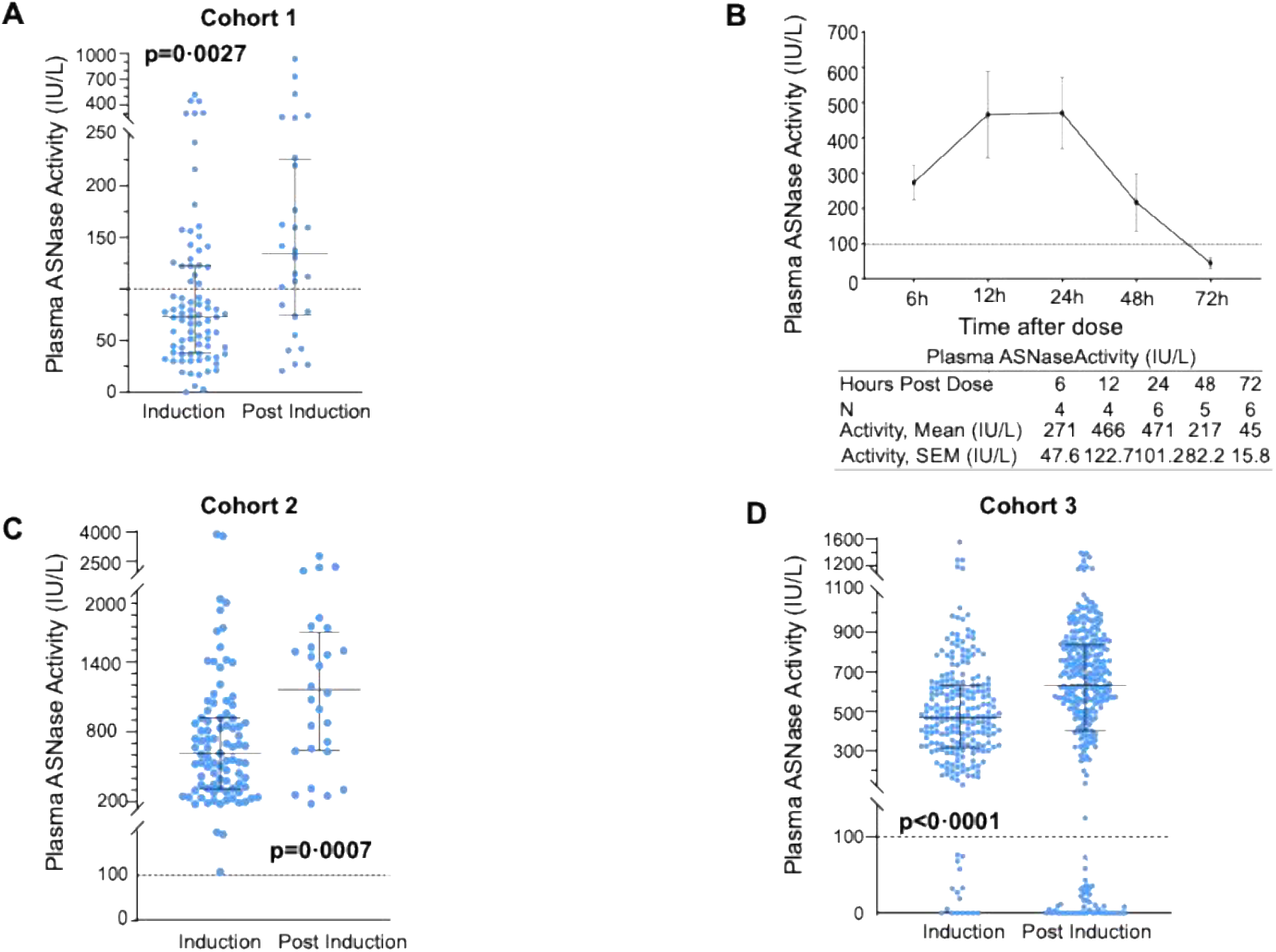
Plasma asparaginase activity measurements across treatment cohorts. (A) Scatter plots of trough plasma asparaginase activity (PAA) measurements in induction and post-induction phases for patients in Cohort 1 (Leucoginase, 10,000 IU/sqm, dosed every 72 hours, intramuscularly). (B) Serial PAA measurements (mean ± standard error) at 6, 12, 24, 48, and 72 hours following a single intramuscular dose of Leucoginase (10,000 IU/sqm) in a subgroup of six patients of Cohort 1. Error bars represent the standard error of the mean (SEM). (C) Scatter plots of trough PAA measurements in Cohort 2 (Leucoginase, 10,000 IU/sqm, dosed every 48 hours, intramuscularly). (D) Scatter plots of trough PAA (12 – 16 days post dose) measurements in Cohort 3, who received PEG-asparaginase (Hamsyl) at 1000 IU/sqm, intramuscularly. In figures A, C, and D, each dot represents PAA measurement of a single plasma sample. The horizontal bars indicate the median and whiskers denote the interquartile range. The dotted horizontal line marks the threshold for satisfactory enzymatic activity (≥100 IU/L). P-values reflect the comparison of PAA between induction and post-induction phases within each cohort.

In all three cohorts, median trough PAA was significantly higher post-induction. Generalised estimating equations (12) were used to model repeated measures of trough PAA in Cohort 3 patients assessing variables potentially influencing these measurements (Supplementary Table S4). Younger age (p=0·0170) and subsequent asparaginase doses (p<0·0001) were significantly associated with higher trough PAA levels. The effect of later doses was partially attenuated by the counteracting influence of asparaginase administration during the delayed intensification treatment phase.

ASNase-associated toxicities, principally clinical hypersensitivity and pancreatitis, varied by drug type (Table 1), and both toxicities necessitated permanent ASNase discontinuation. Clinical hypersensitivity was 2·5 times more frequent in Leucoginase-treated patients (12%; 17/145 patients in Cohorts 1 and 2) than in those treated with Hamsyl (5%; 8/173 patients in Cohort 3) (p=0·0219). Despite the higher number of Leucoginase doses, hypersensitivity was not observed more frequently in Cohort 2 patients (11·6% versus 11·8% in Cohort 1). Pancreatitis occurred nearly four times more frequently with Hamsyl (5%; 9/173) than with Leucoginase (1%; 2/145), although this difference was not statistically significant (p=0·0721). The observed frequency of Hamsyl-associated pancreatitis was comparable to published findings for patients treated with other PEG-EcASNases (16-19). Consistent with published reports (16, 20, 21), patients who developed ASNase-associated pancreatitis were significantly older (median age 11·6 years [IQR, 6·3–15·6]) than patients who did not (median age 5·1 years [IQR, 3·3–8·4]) (p=0·0026).

**TABLE 1.**
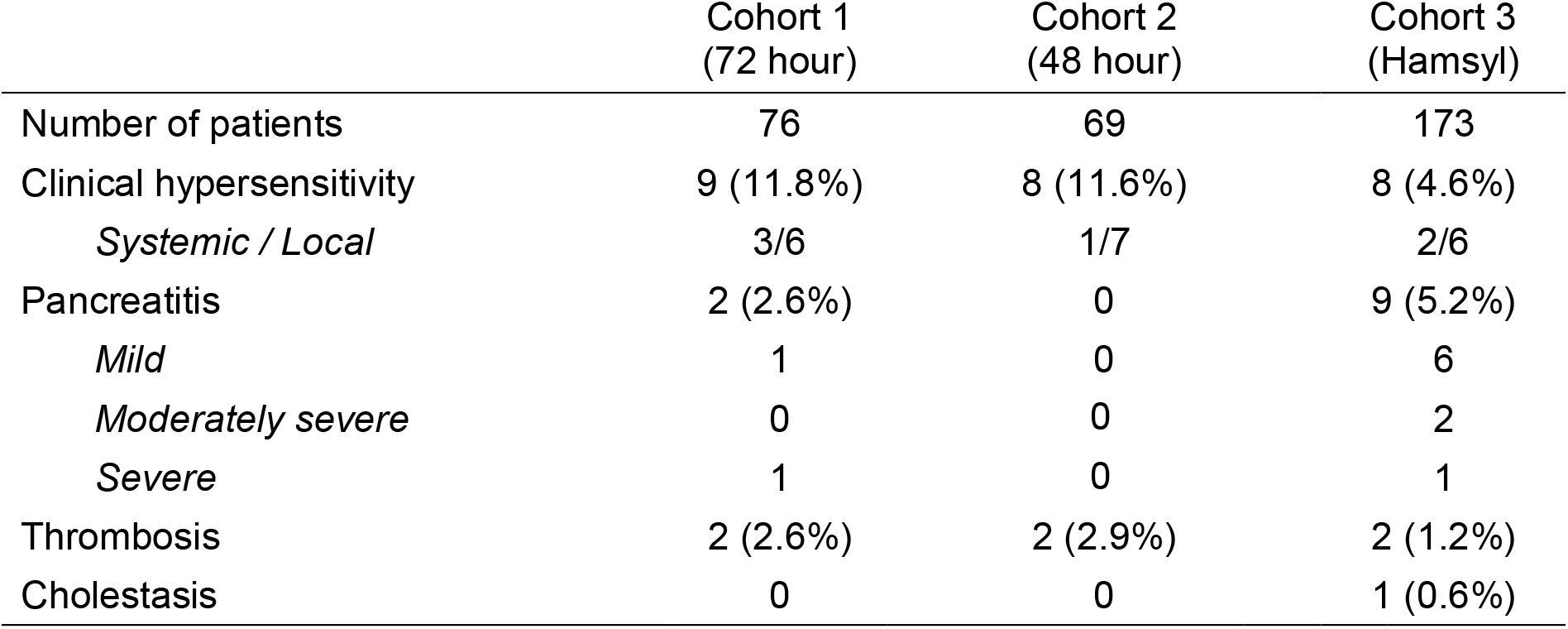
Asparaginase-associated clinical toxicity.

In Cohort 3, paired induction and post-induction samples were available in 139 of 167, allowing identification of the frequency of silent Hamsyl inactivation (Table 2). Sustained adequate PAA was observed in 117 (84%) patients, while seven (5%) had persistent suboptimal activity across induction and post-induction treatment phases. Two patients with suboptimal PAA during induction achieved adequate PAA post-induction. Thirteen patients (9%) with adequate PAA during induction developed suboptimal activity post-induction, suggesting silent inactivation. Patients in Cohort 3 with provisional standard-risk B cell-precursor ALL (SR) received one PEG-EcASNase dose during induction, whereas other risk groups received two doses (Supplementary Table S1). Trough levels were comparable in induction (Supplementary Figure S1). Among the 91 patients receiving two induction doses, median post-induction trough PAA was significantly higher (697 IU/L versus. 484 IU/L, p<0·0001), with similar proportions of subthreshold PAA measurements (4% versus 6%) in induction and post-induction samples (Figure 2, Supplementary Table S5). Of the 48 patients who received a single induction PEG-EcASNase dose (all provisional SR BCP-ALL), trough PAA did not increase significantly post-induction (median, 472 IU/L in induction versus 436 IU/L post-induction). Notably, the proportion of subthreshold PAA measurements was nearly threefold higher post-induction in single-dose recipients (38% vs. 13%, p=0·0015), particularly in those who received additional PEG-EcASNase doses post-induction following reclassification as high-risk (Supplementary Table S6). Consequently, single-dose-induction recipients had approximately eight-fold higher odds (7·8; 95% confidence interval 2·7-20·5) of suboptimal post-induction PAA compared to two-dose-induction recipients (15/48 in one-induction-dose versus 5/91 in two-induction-dose recipients).

**TABLE 2.** Categorisation of Cohort 3 patients based on trough Plasma Asparaginase Activity (PAA) levels at Induction and Post-Induction following Hamsyl PEG-EcASNase Administration.

|  |  | Post Induction |  |  |
| --- | --- | --- | --- | --- |
| PAA |  | ≥100 IU/L | <100 IU/L | Total |
| Induction | ≥100 IU/L | 117 | 13 | 130 |
|  | <100 IU/L | 2 | 7 | 9 |

PAA: Plasma Asparaginase Activity. Adequate: ≥100 IU/L; Subtherapeutic: <100 IU/L.
Note: One patient with transient subtherapeutic activity was excluded. Serial induction and post-induction activity measurements were available in 139 of 167 patients in Cohort 3

**Figure 2.**
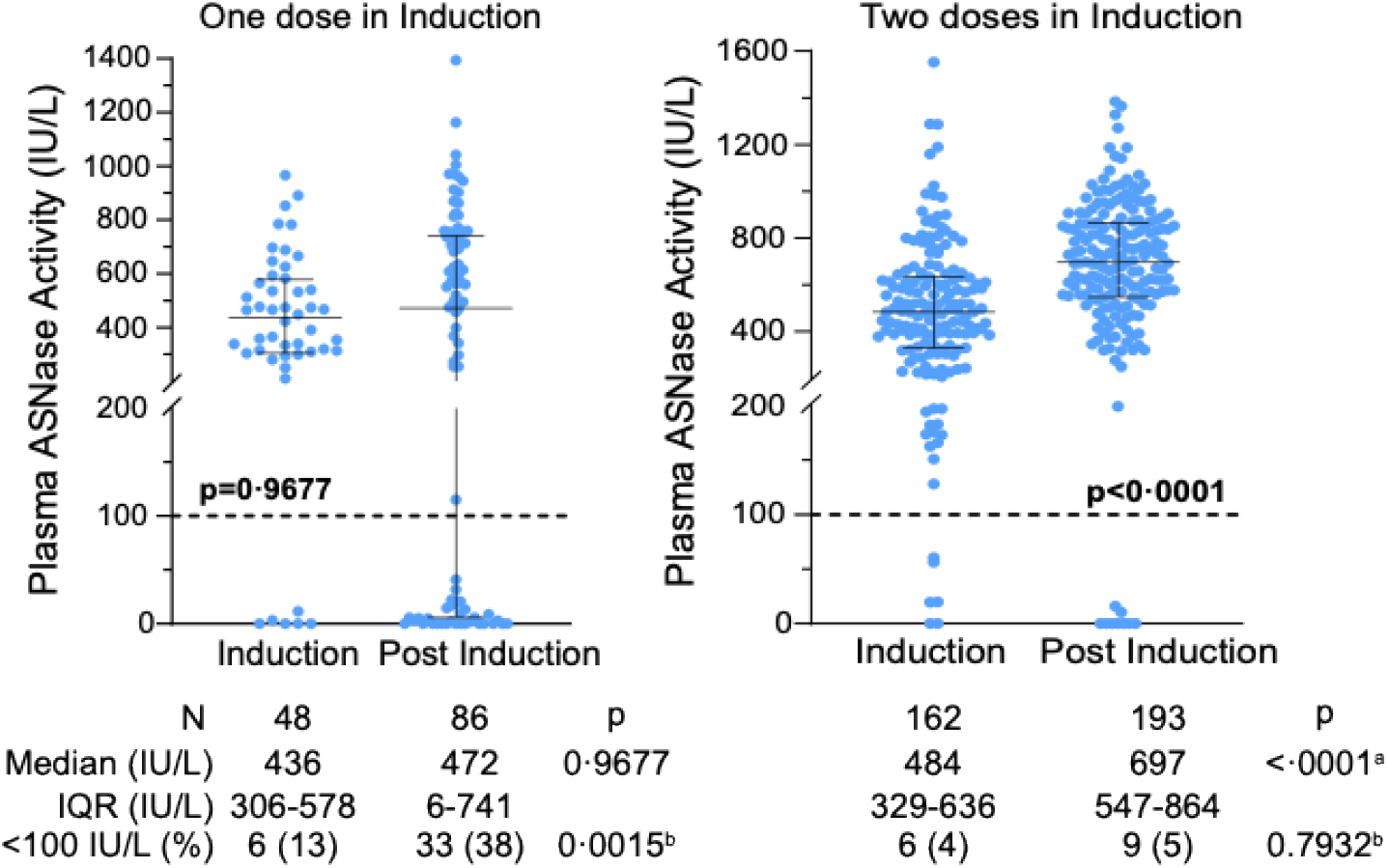
PAA activity at trough timepoints, recorded 12–16 days post administration of PEG-EcASNase across all phases of treatment. (A) in patients receiving one dose or (B) two doses in induction. Each point on the graph represents the measurement of one sample. The central horizontal bars indicate median and whiskers represent the interquartile range. The horizontal dotted line highlights the threshold activity level considered satisfactory (≥ 100 IU/L). P-values reflect the comparison of PAA between induction and post-induction phases within each cohort, ^a^Mann-Whitney test; ^b^Fisher’s Exact Test.

To investigate anti-drug immune responses associated with clinical hypersensitivity and suboptimal asparaginase activity, we measured serial anti-drug antibody (ADA) reactivity in a subset of Cohort 3 patients (Figure 3), including seroreactivity to the intact drug (anti-PEG-EcASNase) and its polyethylene glycol conjugate (anti-PEG). Among 8 patients (P1-P8) diagnosed with ASNase clinical hypersensitivity (Figure 3A), anti-PEG-EcASNase ADA was detected in six (P3-P8), with dual reactivity (to EcASNase and PEG) detected in P7-P8. In P7, dual reactivity was detected at administration of PEG-EcASNase prior to clinical hypersensitivity. In patients P1 and P2, ADA was not detected presumably due to premature sampling (close to the hypersensitivity event in patient P1, likely resulting in assay interference from residual drug) (22) or accelerated clearance of drug-antibody complex (23) (given that ADA and sub-threshold PAA were observed with the Hamsyl dose preceding the hypersensitivity event in patient P2). Trough PAA was inadequate in all patients with clinical hypersensitivity, with sub-optimal activity observed in doses preceding the clinical event. ADA was detected in all four patients (P9-P12) with sustained inadequate PAA across all treatment phases, despite no clinical hypersensitivity. ADA was detected in all 9 patients (P13-P21) with inadequate post-induction PAA, confirming silent inactivation. Three of these patients, P13, P19, P21 showed ADA reactivity with the first asparaginase dose. In these patients, the ADA assay optical density values increased post-induction, suggesting an initial weak non-neutralising immune response that strengthened with repeated drug exposure leading to eventual drug inactivation

**Figure 3.**
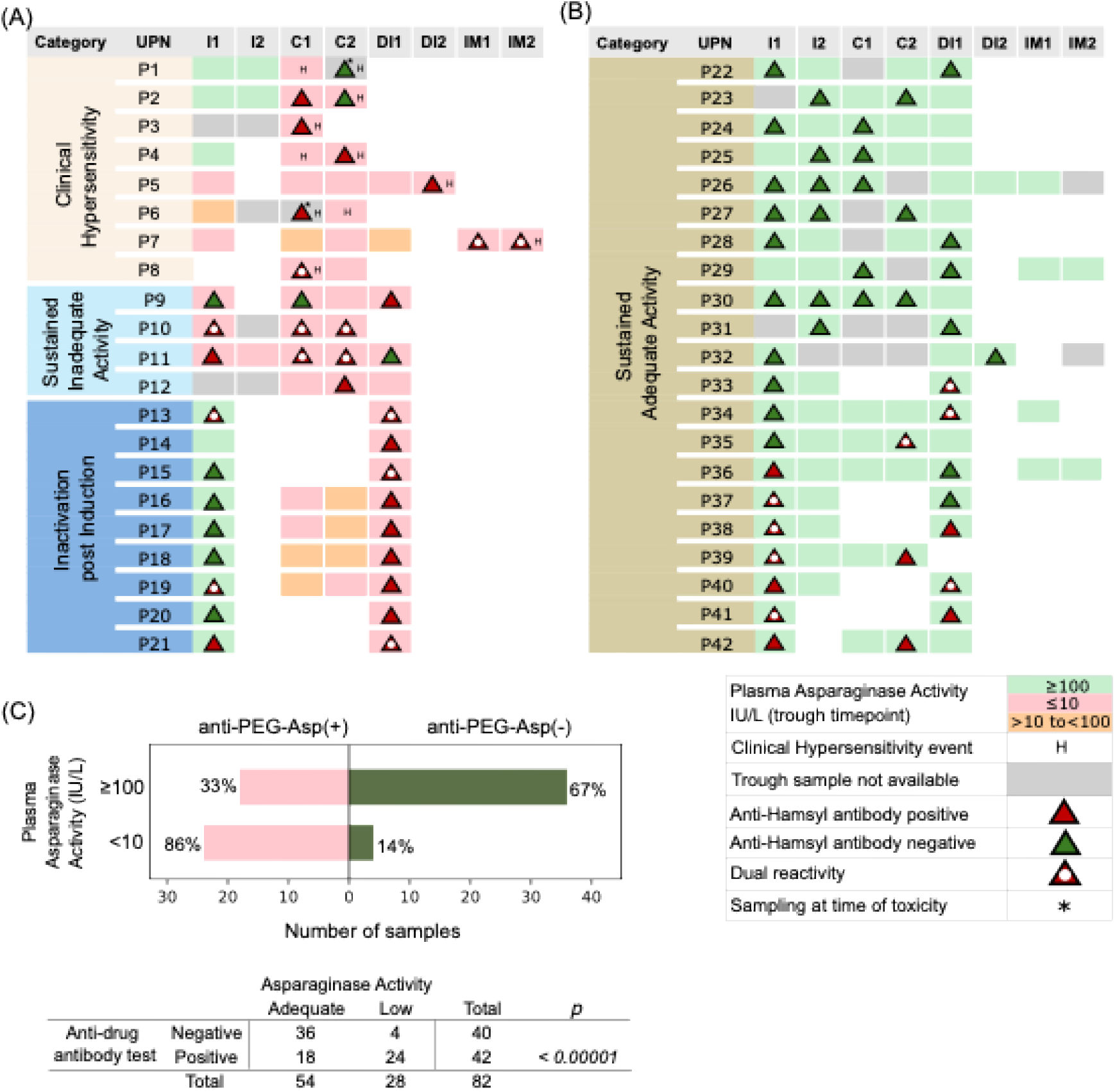
Linking PAA with anti-drug antibody (ADA) reactivity in a subset (n=42) of ALL patients treated with Hamsyl PEG-EcASNAse. Patient categories include those with (A) Clinical hypersensitivity (n=8) and silent inactivation. Silent inactivation includes patients with sustained inactivation (n=4; sky blue) and inactivation observed post induction (n=9; dark blue); and (B) sustained adequate PAA (n=21). I1, I2, C1, C2, DI1, DI2, CM1, CM2 indicate asparaginase doses. In Induction (I1, I2), Consolidation (C1, C2), Delayed Intensification (DI1, DI2) and Capizzi Methotrexate (CM1, CM2) treatment phases; UPN, unique patient number. Further details are available in the Figure Index. Blank cells indicate no clinical sampling at the given timepoint. (C) Horizontal bar chart representing summary of PAA (adequate, ≥ 100 IU/L; inadequate, < 10 IU/L) versus ADA in plasma samples (n=82) from 42 patients. X-axis represents the number of samples demonstrating either ADA reactivity (salmon pink barts) or non-reactivity (olive green bars). The accompanying table highlights the significant association between ADA reactivity and inadequate asparaginase activity (p<0·00001)

Serial testing showed no ADA reactivity in 11/21 (52%) patients with sustained adequate activity (Figure 3B). In three patients (P33–P35), ADA reactivity was detected post-induction with the final or penultimate asparaginase dose. In two patients (P36, P37), transient ADA reactivity was observed, associated with the first asparaginase dose. In five other patients (P38-P42), ADA reactivity was observed at induction and during post induction treatment phases, with assay OD values remaining relatively stable across the phases. Three of these (P38, P39, P41) showed initial dual-reactivity and subsequent EcASNase reactivity alone. Overall, in 82 samples assayed for PAA and ADA reactivity (Figure 3C), inadequate PAA was accompanied by ADA reactivity in 86% samples (24/28), compared to 33% (18/54) of samples with adequate PAA, highlighting the strong association between ADA reactivity and inadequate asparaginase activity (p<0·0001).

## Discussion

This study demonstrates the value of systematic asparaginase monitoring in children with ALL treated on the ICiCle-14 protocol. The 72-hour dosing interval of 10,000 IU/m^2^ per dose for Leucoginase was based on the activity of the Kyowa-Hakko EcASNase. (24) In Cohort 1, 72-hour Leucoginase dosing frequently resulted in subtherapeutic trough activity. Pharmacokinetic profiling confirmed decline in activity below target threshold by 72 hours, and a revised 48-hour schedule adopted in Cohort 2 consistently achieved therapeutic activity without suboptimal levels. A high median trough PAA was observed in Cohort 3 patients treated with Hamsyl (PEG-EcASNase), but ∼9% developed silent inactivation, detectable only through pharmacological monitoring. This was strongly associated with ADA reactivity. Patients receiving a single PEG-EcASNase induction dose were significantly more likely to show inadequate post-induction activity than those receiving two doses, highlighting the importance of adequate induction coverage across risk groups. Toxicity profiles differed by preparation: hypersensitivity was more frequent with Leucoginase, while pancreatitis was more common with Hamsyl, particularly in older children. Though the numbers of patients analysed in this study is small, the results obtained with Hamsyl (PEG-EcASNase) were comparable to those reported for Oncaspar (PEG-EcASNase), both dosed at 1000 iu/m^2^ intramuscularly (12, 25).

Previous studies highlighted substantial concerns regarding the quality of biogeneric asparaginase formulations available in LMICs (3-6), most of which are manufactured and marketed from common sources. (26) Critical concerns such as inconsistent enzymatic activity, purity, and suboptimal therapeutic activity as reported by us, (5) necessitate rigorous clinical validation and are beyond the scope of most hospitals. We discussed our concerns with companies marketing the different formulations. Two, namely VHB Life Sciences and Gennova Biopharmaceuticals, expressed willingness to develop better products and solutions.

In the ICiCLe-ALL-14 trial, SR patients received 4 doses of EcASNase or 1 dose of PEG-EcASNase in induction. This decision was based on the cost and near-unavailability of Oncaspar at ∼£1500 per 3750 IU vial, compared to £15 per 10,000 IU vial for EcASnase, and ready availability of EcASNase biogenerics. The lower dose number was to avoid sensitisation with multiple doses. SR patients treated with 4 doses of EcASNase/1 dose PEG-ASNase were reported to have higher-than-expected relapse rates. (27) In this study, SR patients receiving only a single PEG-EcASNase (1000 iu/m^2^) dose during induction were significantly more likely to have suboptimal post-induction activity compared to those receiving two doses. Increase in ADA levels have been reported when there is a large asparaginase - free interval (14) and PEG-EcASNase administered continuously through induction and post induction, prevents sensitisation without increasing toxicity or affecting outcomes (28). In addition to inadequate asparaginase depletion, reduced early exposure may permit the emergence of immune responses or inadequate asparagine depletion, collectively compromising treatment intensity and increasing the risk of disease recurrence. (2, 6)

ADA reactivity, detected in all patients with persistent or emergent suboptimal activity. Of note, a subset of patients developed ADA reactivity early, with assay signal intensifying on re-exposure, suggesting a progressive immunologic priming that ultimately neutralised drug activity. Detection of ADA in patients with acceptable PAA demonstrates the high antigenicity of the drug and that even patients receiving lymphotoxic and myelosuppressive treatment mount an immune response. There are more than 10 EcASNase and 4 PEG-EcASNases available in the Indian market The presence of ADA without overt hypersensitivity and the association of inadequate activity with relapse suggests that therapeutic drug monitoring (TDM) is essential particularly where the there is insufficient data on the characteristics of the source EcASNase. In the clinic, monitoring ASNase activity is the simplest method to assess the effectiveness of the drug and scheduling. (13) As demonstrated here, real-time pharmacological monitoring to allowed us to refine dosing schedules for EcASNase shorter-acting preparations and introduce a change in dosing of PEG-EcASNAse in induction.

There remains the wide variability of trough PAA after a fixed dose of PEG-EcASNase. As each product is essentially unique, population pharmacokinetic studies can help identify the optimal dose and TDM used to develop individualized dosing strategies. (29, 30) This is required for the Hamsyl product. With the median trough level of 467 IU/L (IQR, 317–630) in induction and higher levels post induction, it is likely that many patients may benefit from a subsequent decreased dose of the drug lowering costs. For patients enrolled into ICiCLe-ALL trials we now recommend PEG-EcASNase as the ASNase of choice and two doses of PEG-EcASNase in induction for all risk categories. Cost remains a major factor. A 1500 IU vial (Hamsyl Junior) is now available at £200 per vial. Assuming 100 patients on a standard 5-dose PEG-EcASNase exposure the total treatment cost is £100,000. TDM in our laboratory costs around £20. If TDM detects inactivation early in 15% of patients allowing timely PEG-EcASNase discontinuation after just two doses instead of five, drug costs will be reduced, both for this group of patients (from £1000 to £400 per patient) and overall (by 9%).

Our findings have significant implications, given ASNase’s essential role in ALL treatment, including its listing on the WHO Essential Medicines for Children and prioritisation by the WHO/St Jude Global Platform for Access to Childhood Cancer, our findings have significant implications. The poor quality of ASNase available in LMICs is likely a major contributor to the inferior survival of children with ALL in LMICs. (31) For patients in LMICs who develop neutralising antibodies, the non-cross reacting Erwinia product is not available. The basic requirement for children with ALL in LMICs is a high quality, affordable PEG-ASNase. The innovator product is unavailable and unaffordable. With multiple biogenerics appearing on the market, our findings reinforce the importance of harmonising pharmacological, immunological, and clinical monitoring across treatment protocols to enable more consistent delivery of this cornerstone of ALL therapy while minimising toxicity.

## Data Availability

All data produced in the present study are available upon reasonable request to the authors

## Acknowledgements

We thank patients and their families for participating in this study; the clinical team who looked after patients and collected data; the hospital laboratory and transfusion medicine departments for supporting the conduct of study, the hospital biorepository for comprehensive management of study samples and Arka Bhowal for laboratory assistance. We thank Gennova Biopharmaceuticals for technical advice in establishing the anti-drug antibody assays. The study was funded in part by a DBT-Wellcome Fellowship (IA/M/12/1/500261) and institute grant from TCS Foundation to VS. Consumable costs were partly defrayed by funding from Gennova Biopharmaceuticals and VHB Life Sciences. The funders did not have a role in the design, conduct, data collection, interpretation of the study or decision to send the paper for publication.

## References

1. Gupta S, Wang C, Raetz EA, Schore R, Salzer WL, Larsen EC, et al. Impact of Asparaginase Discontinuation on Outcome in Childhood Acute Lymphoblastic Leukemia: A Report From the Children’s Oncology Group. J Clin Oncol. 2020;38(17):1897–905.

2. Gottschalk Hojfeldt S, Grell K, Abrahamsson J, Lund B, Vettenranta K, Jonsson OG, et al. Relapse risk following truncation of pegylated asparaginase in childhood acute lymphoblastic leukemia. Blood. 2021;137(17):2373–82.

3. Sankaran H, Sengupta S, Purohit V, Kotagere A, Moulik NR, Prasad M, et al. A comparison of asparaginase activity in generic formulations of E.coli derived L-asparaginase: In-vitro study and retrospective analysis of asparaginase monitoring in pediatric patients with leukemia. Br J Clin Pharmacol. 2020;86(6):1081–8.

4. Cecconello DK, Werlang ICR, Alegretti AP, Hahn MC, de Magalhaes MR, Battistel AP, et al. Monitoring asparaginase activity in middle-income countries. Lancet Oncol. 2018;19(9):1149–50.

5. Sidhu J, Gogoi MP, Agarwal P, Mukherjee T, Saha D, Bose P, et al. Unsatisfactory quality of E. coli asparaginase biogenerics in India: Implications for clinical outcomes in acute lymphoblastic leukaemia. Pediatr Blood Cancer. 2021;68(11):e29046.

6. Michalowski MB, Cecconello DK, Lins MM, Carvalho M, Silva KAS, Cristofani L, et al. Influence of different asparaginase formulations in the prognosis of children with acute lymphocytic leukaemia in Brazil: a multicentre, retrospective controlled study. Br J Haematol. 2021;194(1):168–73.

7. Das N, Banavali S, Bakhshi S, Trehan A, Radhakrishnan V, Seth R, et al. Protocol for ICiCLe-ALL-14 (InPOG-ALL-15-01): a prospective, risk stratified, randomised, multicentre, open label, controlled therapeutic trial for newly diagnosed childhood acute lymphoblastic leukaemia in India. Trials. 2022;23(1):102.

8. Nookala Krishnamurthy M, Narula G, Gandhi K, Awase A, Pandit R, Raut S, et al. Randomized, Parallel Group, Open-Label Bioequivalence Trial of Intramuscular Pegaspargase in Patients With Relapsed Acute Lymphoblastic Leukemia. JCO Glob Oncol. 2020;6:1009–16.

9. Vyas C, Jain S, Kapoor G, Mehta A, Takkar Chugh P. Experience with generic pegylated L-asparaginase in children with acute lymphoblastic leukemia and monitoring of serum asparaginase activity. Pediatr Hematol Oncol. 2018;35(5-6):331–40.

10. Schmiegelow K, Attarbaschi A, Barzilai S, Escherich G, Frandsen TL, Halsey C, et al. Consensus definitions of 14 severe acute toxic effects for childhood lymphoblastic leukaemia treatment: a Delphi consensus. Lancet Oncol. 2016;17(6):e231–e9.

11. Banks PA, Bollen TL, Dervenis C, Gooszen HG, Johnson CD, Sarr MG, et al. Classification of acute pancreatitis--2012: revision of the Atlanta classification and definitions by international consensus. Gut. 2013;62(1):102–11.

12. Sidhu J, Masurekar AN, Gogoi MP, Fong C, Ioannou T, Lodhi T, et al. Activity and toxicity of intramuscular 1000 iu/m(2) polyethylene glycol-E. coli L-asparaginase in the UKALL 2003 and UKALL 2011 clinical trials. Br J Haematol. 2022;198(1):142–50.

13. van der Sluis IM, Vrooman LM, Pieters R, Baruchel A, Escherich G, Goulden N, et al. Consensus expert recommendations for identification and management of asparaginase hypersensitivity and silent inactivation. Haematologica. 2016;101(3):279–85.

14. Tong WH, Pieters R, Kaspers GJ, te Loo DM, Bierings MB, van den Bos C, et al. A prospective study on drug monitoring of PEGasparaginase and Erwinia asparaginase and asparaginase antibodies in pediatric acute lymphoblastic leukemia. Blood. 2014;123(13):2026–33.

15. Woo MH, Hak LJ, Storm MC, Evans WE, Sandlund JT, Rivera GK, et al. Anti-asparaginase antibodies following E. coli asparaginase therapy in pediatric acute lymphoblastic leukemia. Leukemia. 1998;12(10):1527–33.

16. Raja RA, Schmiegelow K, Albertsen BK, Prunsild K, Zeller B, Vaitkeviciene G, et al. Asparaginase-associated pancreatitis in children with acute lymphoblastic leukaemia in the NOPHO ALL2008 protocol. Br J Haematol. 2014;165(1):126–33.

17. Tong WH, Pieters R, de Groot-Kruseman HA, Hop WC, Boos J, Tissing WJ, et al. The toxicity of very prolonged courses of PEGasparaginase or Erwinia asparaginase in relation to asparaginase activity, with a special focus on dyslipidemia. Haematologica. 2014;99(11):1716–21.

18. Chen C, Li J, Chen Y, Gao Q, Li N, Le S. The correlation of asparaginase enzyme activity levels after PEG-asparaginase administration with clinical characteristics and adverse effects in Chinese paediatric patients with acute lymphoblastic leukaemia. Br J Haematol. 2024;205(2):624–33.

19. Kearney SL, Dahlberg SE, Levy DE, Voss SD, Sallan SE, Silverman LB. Clinical course and outcome in children with acute lymphoblastic leukemia and asparaginase-associated pancreatitis. Pediatr Blood Cancer. 2009;53(2):162–7.

20. Liu C, Yang W, Devidas M, Cheng C, Pei D, Smith C, et al. Clinical and Genetic Risk Factors for Acute Pancreatitis in Patients With Acute Lymphoblastic Leukemia. J Clin Oncol. 2016;34(18):2133–40.

21. Oparaji JA, Rose F, Okafor D, Howard A, Turner RL, Orabi AI, et al. Risk Factors for Asparaginase-associated Pancreatitis: A Systematic Review. J Clin Gastroenterol. 2017;51(10):907–13.

22. Myler H, Pedras-Vasconcelos J, Phillips K, Hottenstein CS, Chamberlain P, Devanaryan V, et al. Anti-drug Antibody Validation Testing and Reporting Harmonization. AAPS J. 2021;24(1):4.

23. Gunn GR, 3rd, Sealey DC, Jamali F, Meibohm B, Ghosh S, Shankar G. From the bench to clinical practice: understanding the challenges and uncertainties in immunogenicity testing for biopharmaceuticals. Clin Exp Immunol. 2016;184(2):137–46.

24. Rizzari C, Zucchetti M, Conter V, Diomede L, Bruno A, Gavazzi L, et al. L-asparagine depletion and L-asparaginase activity in children with acute lymphoblastic leukemia receiving i.m. or i.v. Erwinia C. or E. coli L-asparaginase as first exposure. Ann Oncol. 2000;11(2):189–93.

25. Lynggaard LS, Rank CU, Hansen SN, Gottschalk Hojfeldt S, Henriksen LT, Jarvis KB, et al. Asparaginase enzyme activity levels and toxicity in childhood acute lymphoblastic leukemia: a NOPHO ALL2008 study. Blood Adv. 2022;6(1):138–47.

26. Furneaux F, Margottini L. The drug was meant to save children’s lives. Instead, they are dying.. The Bureau of Investrigative Journalism [Internet]. 2023. Available from: https://www.thebureauinvestigates.com/stories/2023-01-25/the-drug-was-meant-to-save-childrens-lives-instead-theyre-dying.

27. Gogoi MP, Das P, Das N, Das S, Narula G, Trehan A, et al. Risk stratified treatment for childhood acute lymphoblastic leukaemia: a multicentre observational study from India. Lancet Reg Health Southeast Asia. 2025;37:100593.

28. van der Sluis IM, Brigitha LJ, Fiocco M, de Groot-Kruseman HA, Bierings M, van den Bos C, et al. Continuous PEGasparaginase Dosing Reduces Hypersensitivity Reactions in Pediatric ALL: A Dutch Childhood Oncology Group ALL11 Randomized Trial. J Clin Oncol. 2024;42(14):1676–86.

29. Boos J, Werber G, Ahlke E, Schulze-Westhoff P, Nowak-Gottl U, Wurthwein G, et al. Monitoring of asparaginase activity and asparagine levels in children on different asparaginase preparations. Eur J Cancer. 1996;32A(9):1544–50.

30. Kloos RQH, Pieters R, Jumelet FMV, de Groot-Kruseman HA, van den Bos C, van der Sluis IM. Individualized Asparaginase Dosing in Childhood Acute Lymphoblastic Leukemia. J Clin Oncol. 2020;38(7):715–24.

31. Krishnan S, Saha V. Global Challenges in Paediatric Acute Lymphoblastic Leukaemia. Acta Haematol. 2025:1–17.

