## Supplemental Tables and Figures for "Biogeneric native and polyethylene glycol-conjugated *E. coli* asparaginases for treating children with acute lymphoblastic leukaemia"

#### Supplementary Information

##### Methods

##### L-Asparaginase treatment

Patients diagnosed with ALL between March 2018 and March 2020 were administered Leucoginase (VHB Life Sciences) native *E.coli* L asparaginase (EcASNase), 10,000 IU/m$^2$/dose intramuscularly. Standard Risk (SR) patients received 4 doses, and others 8 doses in induction. All patients received 4 doses in delayed intensification (DI). High Risk (HR) and T-ALL patients received 8 additional doses during the consolidation phase. Between March 2018 and June 2019, EcASNase was administered at 72-hour dose intervals (Cohort 1), the standard dosing interval for native *E. coli* asparaginase(1). From June 2019 to March 2020 EcASNase dosing interval was modified to once every 48 hours (Cohort 2), resulting in treatment with additional doses of EcASNase during induction (six doses for SR and nine doses for others) and DI (six doses in all risk groups) treatment phases in all patients **(Supplementary Table S1)**.

Patients diagnosed with ALL between August 2020 and March 2023 were treated with PEG-EcASNase (Hamsyl, Gennova Biopharmaceuticals), administered intramuscularly at 1000 IU/m$^2$/dose. SR patients received two doses of Hamsyl (one dose each in induction and DI), Intermediate Risk (IR) patients received three doses (two doses in induction and one dose in DI), and HR and T-ALL patients were administered five doses (two doses in induction and consolidation each and one dose in DI). A subset of patients treated as Very High Risk (VHR), based on persistence of measurable residual disease received two doses of Hamsyl during the interim maintenance and modified DI phases **(Supplementary Table S1)**.

##### Analytical validation of the Asparaginase activity assay

Analytical validation of the plasma ASNase activity assay was performed using pooled fresh frozen plasma (FFP) obtained from the Tata Medical Center Blood Centre, derived from approximately 15 healthy volunteers. Assay validation and performance adhered to established bioanalytical guidelines(2,3).

Assay linearity was established by analysing plasma supplemented with EcASNase concentrations ranging from 5 to 2000 IU/L. Linearity was observed across 10–1500 IU/L with coefficient of variation (%CV) consistently <10% over this range. Precision and accuracy were evaluated by replicate testing in intra-day (8 runs) and inter-day (20 runs) assessments, with %CV consistently <10% in these assessments.

Parallelism was assessed in patient plasma samples representing high, mid, and low ASNase activity levels. Precision (%CV < 10%) was maintained in serial dilutions up to 1:16, accompanied by a mean recovery of ~95%, confirming reliable measurement across dilution ranges. Assay sensitivity was further defined: the lower limit of quantification (LLOQ) was 5 IU/L, the upper limit of quantification (ULOQ) was 1250 IU/L, and the limit of detection (LOD) was 0·98 IU/L.

#### Anti-asparaginase antibody evaluation

Anti-drug antibodies (ADA) against PEG-EcASNase were detected by an indirect enzyme-linked immunosorbent assay (ELISA) adapted from published reports (4,5). Each well of a 96-well ELISA plate was coated with Hamsyl PEG-EcASNAse or lyophilized 5 methoxy-PEG (SinoPEG) and incubated subsequently with test and control plasma samples (1:3200 dilution, 2 hours). Serial dilutions of patient plasma samples were added in triplicate and antibody binding was detected with horseradish peroxidase-conjugated goat anti-human IgG (Sigma), visualised using the tetramethylbenzidine chromogenic substrate. Individual wells were photometrically read at 450 nm on Spectramax M2e (Molecular Devices, LLC). The negative control reference pool was prepared by assaying 45 presumably negative human plasma samples, including from healthy volunteers (n= 23) and from patients with newly diagnosed ALL prior to Hamsyl PEG-EcASNase treatment (n=22). Samples were assessed as positive or negative for ADA based on optical density (OD) readings at 1:3200 dilution. Cutoff for positivity was defined as >1·5 times the third quartile of the negative control, that is >0·123 for the whole molecule (PEG-Asparaginase) and >0·146 for PEG. Dual reactivity was confirmed when antibodies were detected against both the intact drug and its PEG moiety.

#### Results

##### Factors influencing plasma Asparaginase activity

Generalised estimating equations analysis using an exchangeable correlation structure revealed that age, asparaginase dose, and treatment phase significantly influenced plasma asparaginase activity (**Supplementary Table S4**). Age exhibited a negative association with asparaginase activity (-10·95), suggesting decreased activity with increasing age. Higher asparaginase dose number (regression coefficient, 167·9) and delayed intensification treatment phase (regression coefficient 246·8) were each associated with significantly elevated asparaginase activity. However, the interaction between asparaginase dose number and delayed intensification treatment (regression coefficient -126·2) indicated that the individual positive effects of these factors on asparaginase activity were partly offset when combined.

#### References

1. Müller HJ, Beier R, Löning L, Blütters-Sawatzki R, Dörffel W, Maass E, et al. Pharmacokinetics of native Escherichia coli asparaginase (Asparaginase medac) and hypersensitivity reactions in ALL-BFM 95 reinduction treatment. Br J Haematol. 2001 Sep;114(4):794–9.

2. Lanvers C, Vieira Pinheiro JP, Hempel G, Wuerthwein G, Boos J. Analytical validation of a microplate reader-based method for the therapeutic drug monitoring of L-asparaginase in human serum. Anal Biochem. 2002 Oct 1;309(1):117–26.

3. Shah VP, Midha KK, Dighe S, McGilveray IJ, Skelly JP, Yacobi A, et al. Analytical methods validation: bioavailability, bioequivalence and pharmacokinetic studies. Conference report. Eur J Drug Metab Pharmacokinet. 1991;16(4):249–55.

4. Woo MH, Hak LJ, Storm MC, Evans WE, Sandlund JT, Rivera GK, et al. Anti-asparaginase antibodies following E. coli asparaginase therapy in pediatric acute lymphoblastic leukemia. Leukemia. 1998 Oct;12(10):1527–33.

5. Tong WH, Pieters R, Kaspers GJL, te Loo DMWM, Bierings MB, van den Bos C, et al. A prospective study on drug monitoring of PEGasparaginase and Erwinia asparaginase and asparaginase antibodies in pediatric acute lymphoblastic leukemia. Blood. 2014 Mar 27;123(13):2026–33.

**Figure S1.**

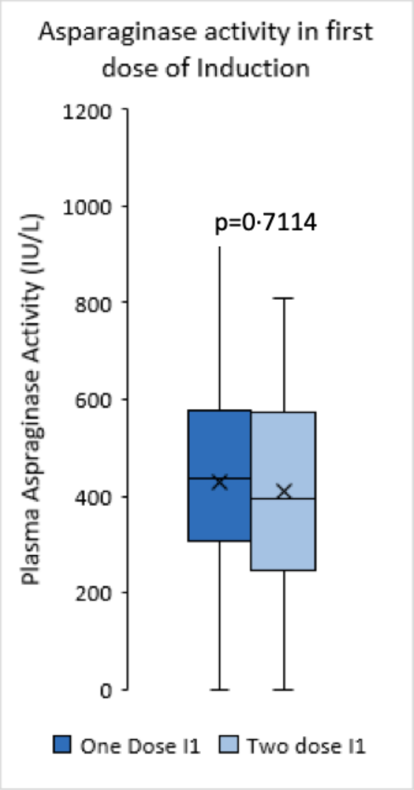


Box-plot representation of plasma asparaginase activity measurements at trough timepoint, recorded 12–16 days post administration of the first dose of Hamsyl PEG-EcASNase in induction, in patients receiving either one dose or two doses of Hamsyl in induction. The central horizontal bars indicate the median, ‘·’ within the boxes indicate mean values and whiskers represent 1·5 times the upper and lower quartiles. P values are calculated using Mann-Whitney U test. I: induction

**TABLE S1**. Asparaginase dosing schedule

| | Number of Doses | | | | |
|---|---|---|---|---|---|
| Phase of Treatment | Ind | Cons | IM | DI | Total |
| **Cohort 1, Leucoginase, q72h** | | | | | |
| Standard Risk | 4 | - | - | 4 | 8 |
| Intermediate Risk | 8 | - | - | 4 | 12 |
| High Risk | 8 | 8 | - | 4 | 20 |
| T-ALL | 8 | 8 | - | 4 | 20 |
| **Cohort 2, Leucoginase, q48h** | | | | | |
| Standard Risk | 6 | - | - | 6 | 12 |
| Intermediate Risk | 9 | - | - | 6 | 15 |
| High Risk | 9 | 8 | - | 6 | 23 |
| T-ALL | 9 | 8 | - | 6 | 23 |
| **Cohort 3, Hamsyl, 1000 IU/m$^2$/dose** | | | | | |
| Standard | 1 | - | - | 1 | 2 |
| Intermediate | 2 | - | - | 1 | 3 |
| High | 2 | 2 | - | 1 | 5 |
| T-ALL | 2 | 2 | - | 1 | 5 |
| Very-High Risk | * | * | 2 | 2 | 4[#] |

Ind = Induction; Cons = Consolidation; IM = Interim Maintenance

DI = Delayed Intensification

q72h, Leucoginase, 10,000 IU/m2/dose, 72-hour dosing interval

q48h, Leucoginase, 10,000 IU/m2/dose, 48-hour dosing interval

* Dosing as per risk assessment

### Four additional doses

**TABLE S2.** Patient characteristics

| | Leucoginase | | Hamsyl |
|---|---|---|---|
| Cohort | Cohort 1 (q72h) | Cohort 2 (q48h) | Cohort 3 |
| Time | Mar 18-Jun 19 | Jun 19-Mar 20 | Aug 20-Mar 23 |
| **N** | 76 | 69 | 173 |
| **Age (years)** | | | |
| Median, IQR | 5·1 (3·8 – 8·8) | 5·5 (3·7 – 8·4) | 5·12 (3·10 – 8·62) |
| **Sex: Male/Female** | 55/21 | 41/28 | 102/71 |
| **WCC** | | | |
| Median, IQR | 17·4 (6·7–71·8) | 12·3 (7·2–33·7) | 16·9 (7·1– 57) |
| **Lineage** | | | |
| B | 59 (78%) | 54 (78%) | 154 (89%)[a] |
| T | 17 (22%) | 15 (22%) | 18 (10%)[b] |
| Mixed phenotype | 0 | 0 | 1 |
| **NCI Risk** | | | |
| Standard Risk | 46 (61%) | 42 (61%) | 91 (53%) |
| High Risk | 30 (39%) | 27 (39%) | 82 (47%) |
| **Cytogenetics (BCP ALL)** | | | |
| Non High Risk | 51 (86%) | 48 (89%) | 137 (79%) |
| High Risk | 2 (3%) | 4 (7%) | 16 (9%) |
| Not available | 6 (10%) | 2 (4%) | 1 (1%) |
| **Risk Group, EoI** | | | |
| BCP ALL Standard Risk | 21 (28%) | 21 (30%) | 36 (21%) |
| BCP ALL Intermediate Risk | 14 (18%) | 10 (14%) | 34 (20%) |
| BCP ALL High Risk | 24 (32%) | 23 (33%) | 83 (48%) |
| T ALL/LL | 17 (22%) | 15 (22%) | 18 (10%) |
| Not applicable* | 0 | 0 | 2 (1%) |

Cohort 1: q72 h, Leucoginase 10000 IU/m$^2$/dose, administered every 72 hours

Cohort 2: q48 h, Leucoginase 10000 IU/m$^2$/dose, administered every 48 hours

IQR, interquartile range; NCI: National Cancer Institute; WCC: White Blood Cell

EoI, End of Induction; EoC, End of Consolidation

[a] Includes five pro B-ALL and one B-cell lymphoma

[b] Includes one T-cell lymphoma

* Patients died before reaching end of Induction

**TABLE S3.** Trough plasma ASNase activity levels in the induction and post-induction phases

| | **Cohort 1 (N=62)** | | | **Cohort 2 (N=57)** | | | **Cohort 3 (N=167)** | | |
|---|---|---|---|---|---|---|---|---|---|
| | Induction | Post Induction | *p* | Induction | Post Induction | *p* | Induction | Post Induction | *p* |
| Samples | 81 | 28 | | 91 | 28 | | 225 | 312 | |
| Median trough activity (IU/L) | 73 | 134 | 0·0027[a] | 617 | 1161 | 0·0007[a] | 467 | 633 | <0·0001[a] |
| Interquartile Range (IU/L) | 38–122 | 74–226 | | 314–918 | 639–1651 | | 317–630 | 403–835 | |
| Mean (IU/L) | 99 | 193 | | 759 | 1185 | | 4849 | 595 | |
| Std. deviation (IU/L) | 98 | 212 | | 638 | 673 | | 262 | 332 | |
| Std. error of mean (IU/L) | 11 | 40 | | 67 | 127 | | 18 | 19 | |
| Proportion below 100 IU/L, n (%) | 56 (69%) | 9 (32%) | 0·0008[b] | 0 | 0 | | 16 (7%) | 49 (16%) | 0·002[b] |

The *p* values are determined using [a] Mann-Whitney U test [b] Chi-Square test.

N, Number of patients tested for plasma Asparaginase Activity

**Table S4.** Generalised estimating equation analysis of variables influencing plasma asparaginase activity in patients treated with Hamsyl PEG-asparaginase

| Covariate | Regression Coefficient | SE | Wald chi-square | Significance |
|---|---|---|---|---|
| *Age (years)* | **-10.95** | 4.59 | 5.693 | **0.0170** |
| *Female Sex* | -39.382 | 38.05 | 1.071 | 0.3007 |
| *T-ALL* | 75.121 | 46.12 | 2.653 | 0.1034 |
| *Risk Group* | | | | |
| BCP-ALL Intermediate Risk | -11.929 | 45.84 | 0.068 | 0.7943 |
| BCP-ALL High Risk / T-ALL | -42.317 | 49.6 | 0.728 | 0.3935 |
| *Treatment Phase* | | | | |
| Consolidation | 59.266 | 111.53 | 0.282 | 0.5954 |
| Interim Maintenance | -489.8 | 455.62 | 1.156 | 0.2823 |
| Delayed Intensification | **246.821** | 100.87 | 5.987 | **0.0144** |
| 2nd Interim Maintenance | -251.249 | 249.19 | 1.017 | 0.3132 |
| *Asparaginase Dose Number* | ***167.891*** | *30.12* | 31.06 | **<0.0001** |
| *Asparaginase Dose Number × Treatment Phase* | | | | |
| × Consolidation | -82.578 | 43.11 | 3.67 | 0.0554 |
| × Interim Maintenance | 15.522 | 86.24 | 0.032 | 0.8580 |
| × Delayed Intensification | **-126.219** | 38.79 | 10.586 | **0.0011** |
| × 2nd Interim Maintenance | -64.55 | 47.42 | 1.853 | 0.1734 |

SE, standard error

Male sex, B cell-precursor ALL (BCP-ALL), Standard Risk BCP-ALL and Induction treatment phase served as reference covariates for Sex, ALL subtype, ALL Risk Group and Treatment Phase respectively

**Table S5.** Serial trough plasma asparaginase activity levels in patients receiving one dose *versus* two doses of Hamsyl PEG-Asparaginase in Induction

| **Treatment Group** | **One dose in induction** | | | | | **Two doses in Induction** | | | | | | |
|---|---|---|---|---|---|---|---|---|---|---|---|---|
| Asparaginase Dose number | 1 | 2 | 3 | 4 | | 1 | 2 | 3 | 4 | 5 | 6 | |
| Protocol Time-point | wk 3 | wk 8 | wk 12 | wk 18/23 | *p* | wk 2 | wk 4 | wk 8 | wk 12 | wk 18/23 | wk 26 | *p* |
| Phase of Treatment | Induction | Consolidation | | DI | | Induction | | Consolidation | | DI | | |
| Total Sample | 48 | 18 | 19 | 46 | | 82 | 80 | 47 | 46 | 79 | 8 | |
| Median (IU/L) | 436 | 68 | 497 | 582 | ns[a] | 396 | 558 | 641 | 648 | 782 | 540 | <0·0001[a] |
| Interquartile Range (IU/L) | 306–578 | 0–514 | 17–814 | 8–757 | | 248–574 | 428–802 | 479–773 | 529–842 | 590–910 | 354–1072 | |
| Proportion <100 IU/L, n (%) | 6 (13) | 9 (50) | 7 (37) | 14 (30) | 0·0090[b] | 5 (6) | 1 (1) | 6 (13) | 2 (4) | 1 (1) | 0 | 0·0213[bc] |

wk, week; DI, Delayed Intensification; ns, not significant

The *p* values are determined using [a] Kruskal Wallis test [b] Fisher's Exact test

**Table S6.** Serial trough plasma Asparaginase activity levels in patients receiving one dose *versus* two doses of PEG-Asparaginase in Induction, categorised by risk group, and post-induction risk continuation or risk escalation

**One dose in Induction**

| Risk group (Initial-Final) | SR-SR | | SR-HR | | | |
|---|---|---|---|---|---|---|
| Phase of treatment-Dose number | Ind-1 | DI-1 | Ind-1 | Con-1 | Con-2 | DI-1 |
| Total sample | 28 | 28 | 20 | 18 | 19 | 18 |
| Median (IU/L) | 504 | 590 | 337 | 69 | 497 | 576 |
| Interquartile Range (IU/L) | 336 – 683 | 90 – 836 | 228 – 468 | 0 – 514 | 17 – 814 | 3 – 741 |
| Proportion <100 IU/L, n (%) | 2 (7) | 7 (25) | 4 (20) | 9 (50) | 7 (37) | 7 (39) |

**Two doses in Induction**

| Risk group (Initial-Final) | IR-IR | | | IR-HR and HR/T | | | | | |
|---|---|---|---|---|---|---|---|---|---|
| Phase of treatment-Dose number | Ind-1 | Ind-2 | DI-1 | Ind-1 | Ind-2 | Con-1 | Con-2 | DI-1 | DI-2 |
| Total sample | 28 | 28 | 30 | 54 | 52 | 47 | 46 | 49 | 8 |
| Median (IU/L) | 392 | 668 | 733 | 412 | 505 | 641 | 648 | 791 | 540 |
| Interquartile Range (IU/L) | 230 – 601 | 453 – 871 | 585 – 907 | 277 – 569 | 416 – 734 | 479 – 773 | 529 – 842 | 595 – 929 | 354 – 1072 |
| Proportion <100 IU/L, n (%) | 1 (4) | 0 | 0 | 4 (7) | 1 (2) | 6 (13) | 2 (4) | 1 (2) | 0 |

SR, standard risk; IR, intermediate risk; HR, high risk; Ind, induction; Con, consolidation; DI, delayed intensification
SR-SR, Initial (Induction) and Final (Post-Induction) Risk as SR
IR-IR, Initial (Induction) and Final (Post-Induction) Risk as IR
SR-HR and IR-HR, Initial (Induction) Risk as SR and IR respectively, and Final (Post-Induction) Risk as HR
HR/T, patients classified as HR and patients with T-ALL, treated accordingly in Induction and post-Induction phases
